# Specific Symptom Profiles are Associated with Distinct Changes in Amygdala Connectivity in Youth

**DOI:** 10.1101/19010165

**Authors:** Joel Stoddard, Katharina Kircanski, Simone P. Haller, Kendra E. Hinton, Banafsheh Sharif-Askary, Melissa A. Brotman

## Abstract

Complex clinical presentations are common in youth seeking mental health treatment, complicating attempts to identify specific biological underpinnings to guide precision psychiatry. We defined four classes of such youth based on their symptom profiles and identified unique patterns of amygdala functional connectivity in each class.

Subjects were 215 youth who varied along major symptom dimensions commonly associated with pediatric affective psychopathology: depression, irritability, anxiety, and attention-deficit/hyperactivity (ADHD). We used latent profile analysis to identify classes of symptom patterns. Functional MRI data were obtained while subjects completed a gender identification task of face-emotions that varied in emotion type and intensity. We used generalized psychophysiological interaction analysis to examine associations between the probability of being in each symptom class and amygdala functional connectivity.

The likelihood of being in the class with high parent-reported irritability and ADHD symptoms was associated with amygdala connectivity to the insula and superior temporal gyrus while processing high-intensity angry and fearful faces; to the precuneus while processing intensity across emotions; and to the ventrolateral prefrontal cortex across all task conditions. The likelihood of being in the class with high anxious and depressive symptoms was negatively associated with amygdala-thalamic connectivity across task conditions.

This is the first study to identify distinct associations between symptom profile classes and amygdala connectivity in a transdiagnostic sample of youth. These neural correlates provide external validity to latent classes derived from symptom clusters. This is an essential first step toward identifying the biological basis of common transdiagnostic symptom presentations in youth.

## Introduction

Youth presenting for mental health care often have multiple symptoms and diagnoses, complicating the prescription of targeted treatment interventions. One way to address this complexity is to use person-centered approaches to understand the pathophysiology underlying different patterns of co-occurring symptoms (Friston *et al*, 2017; Insel, 2009). With the development of mixture models for detecting subpopulations, it is now possible to classify individuals by naturally-occurring symptom patterns, or profiles (Goodman, 1974; Hagenaars and McCutcheon, 2002). Recently, using dimensions of psychopathology, we classified youth empirically into broad profile classes (Kircanski *et al*, 2017). However, to improve diagnosis and treatment, it is essential to validate these classes such as by identifying their neural basis. Here, we take this approach by assessing associations between profile classes and amygdala connectivity while processing face-emotions.

In prior work, we used a type of mixture model, latent profile analysis (LPA), and parent and child symptom report to classify 509 youth participating in psychiatric research on affective psychopathology. The analysis generated five profile classes, including groups, which were characterized by 1) low levels of all symptoms (52% of sample); 2) moderate anxious and mild depressive symptoms (17%); 3) high parent-reported irritability and attention-deficit/hyperactivity symptoms (16%); 4) high irritability and mixed comorbid symptoms (10%); and 5) high levels of all symptoms (5%). These five classes had differential, expected associations with demographics and severity. However, an important limitation of this and other symptom-based classifications is its reliance on measures without a known biological basis.

An understanding of the neural mechanisms mediating different symptom patterns would advance diagnosis, support treatment development, and facilitate precision medicine initiatives (Insel, 2009). Indeed, recent examinations of data-rich brain connectivity images reveal meaningful associations between brain function and such data-driven, symptom-based classifications. These efforts are already showing promise for elucidating biologically-driven subtypes of mental illnesses (Drysdale *et al*, 2017; Karalunas *et al*, 2014; Satterthwaite *et al*, 2016; Tamminga *et al*, 2014; Van Dam *et al*, 2017) and guiding treatment selection (Doehrmann *et al*, 2013; Dunlop *et al*, 2017).

The goal of this study is to validate our empirical profile classes by examining their unique associations with amygdala-based functional connectivity during face-emotion processing. Amygdala networks underlying emotion regulation and threat processing are implicated in the affective symptoms that characterize our target population (Bebko *et al*, 2015; Hafeman *et al*, 2017; Jalbrzikowski *et al*; Karalunas *et al*, 2014; Kujawa *et al*, 2016; Stoddard *et al*, 2017). Indeed, amygdala connectivity during face-emotion processing may be a more reliable measure than either activation or behavior (White *et al*, 2016). Thus, task-based amygdala connectivity is a promising candidate for fMRI-based biomarker studies of affective psychopathology.

Here, we examine data from 215 youth who completed a commonly-used implicit face-emotion processing fMRI task. We used a data-driven method to classify individuals by symptom profile and assessed associations between task-related amygdala connectivity and membership in a symptom profile class. We expected to find associations between class membership and connectivity within amygdala-based circuitry previously implicated in face-emotion processing and pediatric affective psychopathology, in particular amygdala-prefrontal (Hafeman *et al*, 2017; Jalbrzikowski *et al*; Stoddard *et al*, 2014; White *et al*, 2017) and amygdala-insular (Bebko *et al*, 2015; Karalunas *et al*, 2014; White *et al*, 2017) circuitry have been found to associated with specific aspects of affective psychopathology in youth. We also used a whole brain, exploratory approach to examine associations between brain activity and membership in a symptom profile class (Friston, 2011).

## Methods

### Participants and Measures

The sample included 215 youth (45.6% female) aged 7 through 18 years [M (SD) =13.6 (2.7) years] who participated in research at the NIMH between 2012 and 2015 and completed an fMRI implicit emotion processing task. Participants were initially recruited into the following groups based on primary diagnosis: anxiety disorder (ANX, n=42, generalized anxiety disorder, social anxiety disorder, or separation anxiety disorder), disruptive mood dysregulation disorder (DMDD, n=44), attention deficit/hyperactivity disorder (ADHD, n=32), bipolar I or II disorder (BD, n=16). The sample also included subjects who had a first-degree relative with bipolar disorder (“at risk” AR, n=35), and those with no psychopathology (healthy volunteers, HV, n=46). Of the 215 participants, 185 were also included in our original LPA of 509 youth (Kircanski *et al*, 2017); however, that study did not include neuroimaging data. Additionally, 113 of the 215 subjects contributed imaging data to a study of the effects of irritability and anxiety on amygdala connectivity during implicit face-emotion processing (Stoddard *et al*, 2017). Thus, the current study contains data in an additional 112 subjects, beyond those appearing in this prior imaging paper. Comparisons among samples and results of this study, Kircanski *et al*, (2017) and Stoddard *et al*, (2017) are detailed in Supplementary Materials and Methods.

The clinical assessment and behavioral measures are as described by Kircanski *et al*, (2017). Exclusion criteria for all groups were unstable medical conditions, autism spectrum disorder, substance use within 2 months prior to scan, head trauma, and intelligence quotient (IQ) <70. Individuals who did not complete a scan or had poor task performance (<65% accuracy) were not included. Ten otherwise eligible youth completed a scan but were excluded due to poor scan quality or processing errors (Table S1).

The study was approved by the NIMH Institutional Review Board. Written informed consent was obtained from parents and assent from children. Families were paid for participation.

To maximize accrual, relatives were allowed in this study, resulting in 18 sibling pairs and 4 sibling triads. To ensure that results were not due to sibship, results were confirmed in analyses that included only one randomly chosen individual per sibling group (n=189; Table S2).

The sum of items of each of the parent- and self-report Affective Reactivity Index (ARI) (Stringaris *et al*, 2012) provided two measures of irritability. The sum of items of each of the parent- and self-report Screen for Child Anxiety Related Disorders (SCARED) (Birmaher *et al*, 1999) provided two measures of anxiety. The sum of items of the self-report Children’s Depression Inventory (CDI) (Kovacs, 1978) measured depressive symptoms. The ADHD Index T-score on the Conner’s Parent Rating Scale (CPRS) (Conners, 1989) measured ADHD symptoms. Differing from Kircanski et al, (2017), we used the ADHD Index T-score rather than the sum of ADHD subscale items to represent ADHD symptoms.

Relationships among symptom reports, age, intelligence, gender, and motion during scanning are presented in Table 1.

**Table 1:**
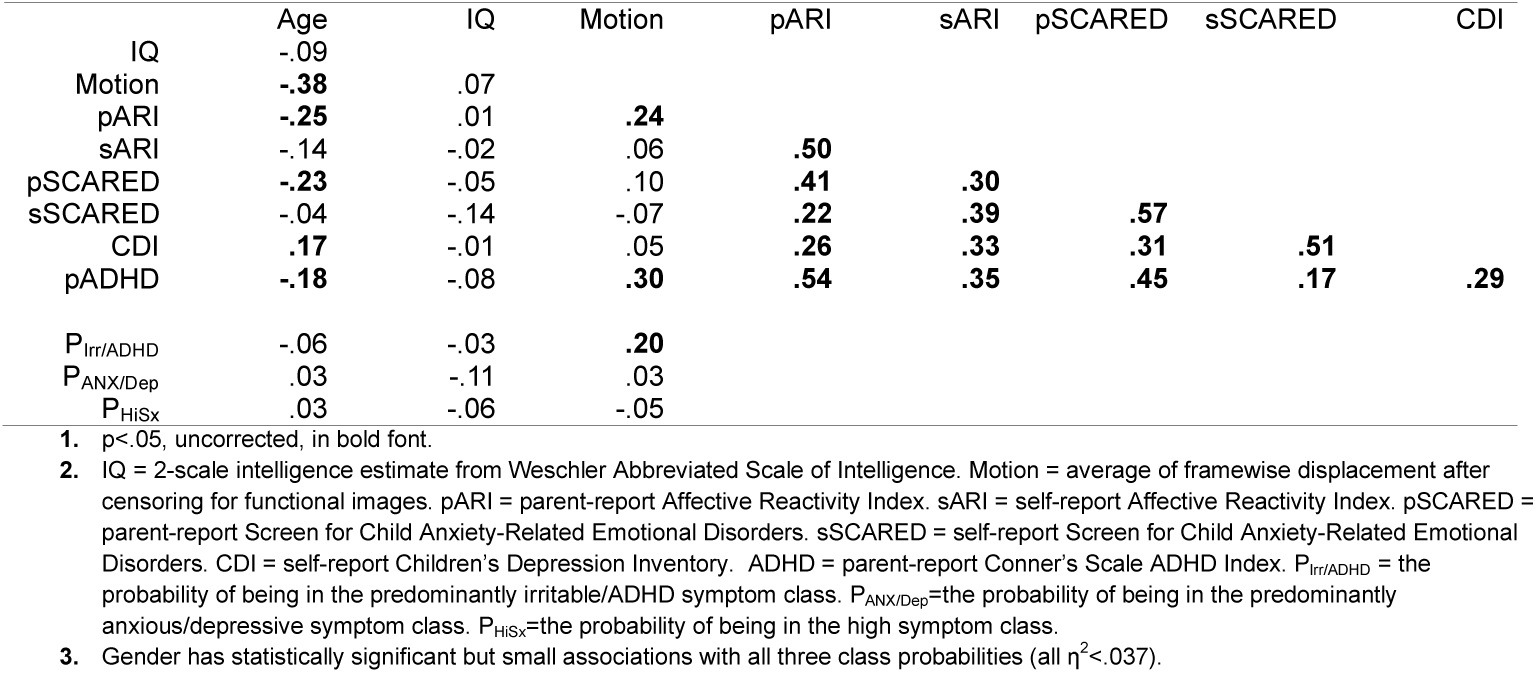
Pearson’ s Correlations^1^ among Participant Characteristics^2,3^.

|  | Age | IQ | Motion | pARI | sARI | pSCARED | sSCARED | CDI |
| --- | --- | --- | --- | --- | --- | --- | --- | --- |
| IQ | -.09 |  |  |  |  |  |  |  |
| Motion | **-.38** | .07 |  |  |  |  |  |  |
| pARI | **-.25** | .01 | **.24** |  |  |  |  |  |
| sARI | -.14 | -.02 | .06 | **.50** |  |  |  |  |
| pSCARED | **-.23** | -.05 | .10 | **.41** | **.30** |  |  |  |
| sSCARED | -.04 | -.14 | -.07 | **.22** | **.39** | **.57** |  |  |
| CDI | **.17** | -.01 | .05 | **.26** | **.33** | **.31** | **.51** |  |
| pADHD | **-.18** | -.08 | **.30** | **.54** | **.35** | **.45** | **.17** | **.29** |
| P<sub>Irr/ADHD</sub> | -.06 | -.03 | **.20** |  |  |  |  |  |
| P<sub>ANX/Dep</sub> | .03 | -.11 | .03 |  |  |  |  |  |
| P<sub>HiSx</sub> | .03 | -.06 | -.05 |  |  |  |  |  |

1. $p < .05$ , uncorrected, in bold font.
2. IQ = 2-scale intelligence estimate from Weschler Abbreviated Scale of Intelligence. Motion = average of framewise displacement after censoring for functional images. pARI = parent-report Affective Reactivity Index. sARI = self-report Affective Reactivity Index. pSCARED = parent-report Screen for Child Anxiety-Related Emotional Disorders. sSCARED = self-report Screen for Child Anxiety-Related Emotional Disorders. CDI = self-report Children's Depression Inventory. ADHD = parent-report Conner's Scale ADHD Index. P<sub>Irr/ADHD</sub> = the probability of being in the predominantly irritable/ADHD symptom class. P<sub>ANX/Dep</sub> = the probability of being in the predominantly anxious/depressive symptom class. P<sub>HiSx</sub> = the probability of being in the high symptom class.
3. Gender has statistically significant but small associations with all three class probabilities (all $\eta^2 < .037$ ).

### Latent Profile Analysis

As in our original LPA (Kircanski *et al*, 2017), manifest variables in the LPA were separate parent- and child-reported ARI and SCARED scores, but only self-reported CDI and parent-reported CPRS scores. To be included in the LPA, a measure must have been collected within 60 days of scanning. Up to 15% of items could be missing on any measure; missing items were imputed from remaining items. Of 215 participants, 129 had a complete set of six measures, 49 had five, 16 had four, and 21 had three or fewer. There were no relationships between missing a measure and age, intelligence, or gender (p’s>0.45). Missing a measure was not related to ultimate class assignment (p=0.49). Given the wide range in age, standardized residuals from each measure regressed on age were entered into the LPA.

We used Mplus Version 7.4 (Muthén and Muthén, 1998) to derive latent classes. The parameters were the same as in the original analysis (Kircanski *et al*, 2017), except the number of classes. As described in the introduction, the original analysis yielded a 5-class solution. Here, a 4-class solution (entropy=0.78) resulted in the most similar set of categories, in terms of symptom profile and class assignment in the overlapping samples (Supplementary Materials and Methods). The four classes were characterized by high levels of all symptoms (Hi Sx), low levels of all symptoms (Lo Sx), moderate irritability and ADHD symptoms (Irr/ADHD), and moderate anxiety and depressive symptoms (Anx/Dep) (Figure 1). Differences in the number of classes reflect differences in the samples included in the current, vs. the original, LPA (Supplementary Materials and Methods). Largely because the current study required that participants be suitable for fMRI, it had 294 fewer subjects than the original study. Of the 185 participants included in both Kircanski *et al*. (2017) and the current study, 70% were assigned to the corresponding group in both studies (Supplementary Materials and Methods). The original analysis also contained a “moderate irritability and mixed comorbid symptoms,” which in this study was evenly distributed among the current Hi Sx and Irr/ADHD groups.

**Figure 1.**
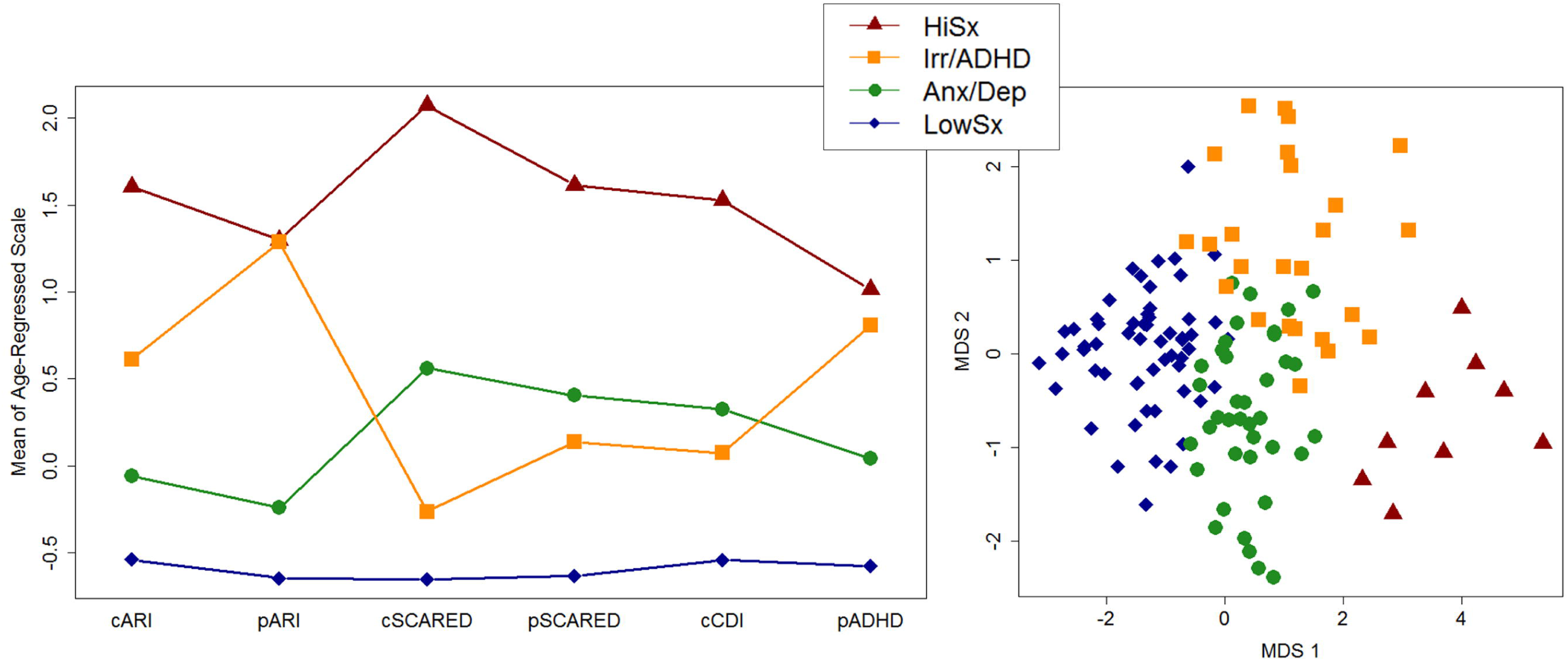
Two views of clinical data for the LPA four-class solution. The left panel is a profile plot with a line for each class. They are low symptoms on all measures (LoSx), relatively higher parent report of irritability and ADHD symptoms (Irr/ADHD), relatively higher anxiety and depression symptoms (ANX/Dep), and high symptoms on all measures (HiSx). We regressed each symptom measure (x-axis) on age across the whole sample and then entered the resultant standardized residuals into LPA. For each LPA class, the mean of these standardized residuals are plotted on the y-axis. Thus, the y-axis is a common scale for all measures, with a score of zero representing the mean score of each measure across the entire sample. The lines for each class may be interpreted as the characteristic symptom level on each measure for members of the class. The right panel represents individuals with a complete set of measures (n=129) plotted by symptom dimension. We used multidimensional scaling (MDS) to collapse all six clinical measures into two components (MDS1 and MDS2) for visualization. As expected, the first component (MDS1) separates individuals by overall degree of symptomatology, i.e. general severity. Plot points have colors and shapes by LPA class. Individuals assigned to each LPA class cluster together by MDS symptom dimensions, suggesting agreement between MDS and LPA methods. However, the proximity of individuals at class-cluster borders suggests individuals assigned to one class may be similar to individuals assigned to another class in symptoms. This supports our analytic approach of using class probability rather than class assignment to preserve information on uncertainty of class assignment.

### Probability of Class Assignment

Individuals differ in the in their goodness of fit to the classes identified by the group analysis (Figure 1). Mixture modeling, like the LPA used here, can account for this individual variation by providing the probability of an individual’s assignment to each class. Using the probability of class assignment as a predictor (instead of class assignment per se) avoids the assumption that there are no differences between individuals within a class (Whalen, 2017). Also, by using the probability of class assignment in our analysis, we can weigh the influence of each person on the results, making the analysis dimensional and hence more powerful. Thus, we used the LPA probability of class assignment in our analyses. We denote this as P_class_ overall or, for the probability of being in a specific class, by P and a subscript corresponding to the class, e.g., P_LoSx_.

### fMRI Task

During fMRI, participants labelled the gender of happy, angry, and fearful face-emotion pictures from 10 actors (Ekman and Friesen, 1976). Stimuli with varying intensity of expression (50%, 100% and 150%) were created by morphing emotional faces with neutral faces. Face stimuli were presented in random order for 2s followed by a jittered fixation cross (mean duration1.4s, range 0.5-6s). Trials were divided into 3 blocks with a total of 30 trials for each emotion by intensity condition and 90 neutral face-emotion trials (Stoddard *et al*, 2017).

### Imaging Procedures

Magnetic resonance images (MRI) were acquired on a General Electric 3.0 Tesla scanner with a 32-channel head coil. Blood oxygen-level dependent (BOLD) signal was measured by echoplanar imaging (EPI) at 2.5 × 2.5 × 3.0mm voxel resolution resampled to 2.5mm isotropic. One mm isotropic T1 images for anatomic registration and nonlinear normalization were acquired with a magnetization-prepared rapid acquisition gradient echo sequence (MPRAGE; flip angle = 7 degrees, minimum full echo time, inversion time = 425ms, acquisition voxel size = 1 mm isotropic). All images were visually inspected for acquisition artifact and proper registration. This work used the computational resources of the NIH HPC Biowulf cluster (http://hpc.nih.gov).

Images were processed using FreeSurfer (Ségonne *et al*, 2004) and Analysis of Functional Neuroimages (AFNI) (Cox, 1996) software. MPRAGE images were uniformity corrected and skull stripped with FreeSurfer (Ségonne *et al*, 2004) and normalized via nonlinear registration to AFNI’s TT_N27 Taliarach-space template. EPI images were processed by removing the first four pre-magnetization volumes, limiting each voxel’s BOLD signal to four standard deviations from the mean trend of its time series, correcting for slice timing, affine volume to volume and volume to anatomic registration, smoothing using a 5 mm FWHM Gaussian kernel, and scaling to a mean of 100.

Processed, scaled EPI were then entered into a general linear model (GLM) with the following parameters: a cubic detrending polynomial, a regressor for each of six translational and rotational motion parameters, physiologic noise regressors (one each for the first 3 principle components of all voxels timeseries in an eroded white matter and lateral ventricle mask (Behzadi *et al*, 2007), a 2 second block-GAM convolved regressor for each emotion by intensity stimulus class, neutral faces, and inaccurate trials. The residuals from this GLM were then entered into a generalized psychophysiologic interaction analysis (gPPI; McLaren *et al*, 2012).

For amygdala connectivity, we averaged timeseries from processed, scaled EPI signal across voxels within each amygdala defined by the AFNI DKD_MPM atlas. For each subject, each of the average amygdala timeseries was partitioned by task condition for gPPI regressors (Stoddard *et al*, 2017). For each amygdala, the residuals from the single subject GLM were entered into a, second GLM with the regressors from the first model in addition to gPPI regressors and a regressor for the average amygdala timeseries. In the second GLM only, EPI volumes were censored from this GLM regression by three criteria: 1) motion shift, defined as movement exceeding a Euclidean distance of 1 mm from its preceding volume, 2) volumes immediately preceding such a motion shift, and 3) volumes with >10% voxels at outlying time points. Subjects were excluded if more than 15% of their volumes were censored or their mean censored volume to volume Euclidean displacement was more than or equal to 0.2mm (Table S1). Parameter estimates for gPPI regressors for each emotion by intensity represent the amplitude of connectivity and were entered into second-level group analyses.

### Analyses

Analyses were conducted in AFNI or R (R Core Team, 2014). All omnibus analyses used a linear mixed-effects model-based ANOVA implemented in 3dLME (Chen *et al*, 2014) for images or in the lme function in R package nlme (Singmann *et al*, 2015) for behavior or *post hoc* analyses of imaging results.

The ANOVA tested the fully interactive effects of each Probability of Class Assignment (P_class_) with Emotion and Intensity, except that no term could include an interaction between two or more P_class_. Age, Motion (mean Euclidean distance), and Gender were covariates. Emotion and Intensity were modeled as within-subjects factors with three levels (Emotion: Happy, Angry, Fearful; Intensity: 50%, 100%, 150%). Age and Motion were between-subjects, continuous variables centered at their mean across the sample. Gender was a between-subjects factor. Subject was a random factor. Because P_LoSx_ was anticorrelated to each of P_HiSx_, P_Irr/ADHD_, and P_Anx/Dep_, it was not included to prevent variance inflation by multicollinearity.

For behavioral analyses, percent correct gender identification (accuracy) and mean reaction time for trials with correct gender identification were dependent variables. For imaging analyses, amygdala connectivity during each emotion by intensity condition, relative to fixation, were dependent variables. Results from these models may be interpreted as the unique association between P_class_ by Emotion and Intensity to behavior or amygdala connectivity.

The imaging analysis was conducted across a whole brain mask, including only voxels where data existed for at least 90% of subjects. The primary voxelwise p-value threshold was set at 0.005. Multiple testing was corrected to α = 0.05 over the whole experiment, Bonferroni corrected for two group tests. This was done via Monte Carlo simulation with a Gaussian plus exponential term spatial autocorrelation function to estimate smoothness (3dClustSim with –acf option; α threshold= 0.05/2= 0.025) (Cox *et al*, 2016). The resultant cluster size threshold was 100 contiguous voxels, each sharing at least one edge with its neighbor. Clusters are reported with size (k) and coordinates of the peak of a cluster with RAI convention in Taliarach space. For further descriptive analysis of each functional ROI, mean values of connectivity at each emotion by intensity were extracted via AFNI’s 3dROIstat.

For *post hoc* analyses, we fit mixed effects models using the same formula as fMRI group analysis. We used general linear tests (Pillai’s Trace) of specific contrasts or fixed effects, adjusting for all others in R package phia (De Rosario-Martinez, 2013). These were Holm-Bonferroni corrected for multiple comparisons. To minimize Type I error, significance tests were re-run without influential participants. Influential participants were defined by a Cook’s Distance > 0.02, a threshold based on the sample size and number of model parameters (Fox, 1991). Any result no longer significant when influential subjects were removed was not reported.

## Results

All results are of associations between behavior or brain function and the degree to which an individual is associated with an LPA class (Probability of Class Assignment, P_class,_ Figure 1). P_Anx/Dep_ > 0.1 for n=100; P_Irr/ADHD_ > 0.1 for n=61; P_HiSx_ > 0.1 for n=20.

### Behavior

There were no associations between any P_class_ and reaction time or accuracy.

### Unique Associations of Amygdala Connectivity to Symptom Classes

For regions showing unique associations between each symptom class and amygdala connectivity, see Table 2 and Figure 2. Classes were associated with task modulated amygdala connectivity in six regions.

**Table 2:** Brain Regions with Unique Associations between Amygdala Connectivity and Symptom Class.

| **Class** | **Seed** | **Connectivity Target Region<sup>1</sup>** | **Size (k)** | **Peak (RAI xyz)** | **Significant Term<sup>2</sup>** | **Comparison** | **Post hoc analysis<sup>3</sup>** |
| --- | --- | --- | --- | --- | --- | --- | --- |
| Irr/ADHD | L amy. | L STG, insula | 126 | 51.2, 1.2, 6.2 | $P_{\text{Irr/ADHD}} \times E \times I$ | $F(4,1688)=6.96$,<br>$p<.001$ | A150, F150 < H150 |
| Irr/ADHD | R amy. | B precuneus | 114 | 6.2, 71.2, 28.8 | $P_{\text{Irr/ADHD}} \times I$ | $F(1,1688)=9.8$,<br>$p<.001$ | 50 > 100, 150 |
| Irr/ADHD | L amy. | L IOFC | 125 | 43.8, -23.8, -3.8 | $P_{\text{Irr/ADHD}}$ | $F(1,208)=21.5$,<br>$p<.001$ | Negative association ($r=-.31$)<br>between $P_{\text{Irr/ADHD}}$ and connectivity |
| Anx/Dep | L amy. | R thalamus | 101 | -13.8, 13.8, 3.8 | $P_{\text{Anx/Dep}}$ | $F(1,208)=22.7$,<br>$p<.001$ | Negative association ($r=-.28$)<br>between $P_{\text{Anx/Dep}}$ and connectivity |
| Hi Sx | R amy. | L lingual gy.,<br>occipital gy. | 123 | 18.8, 91.2, -8.8 | $P_{\text{HiSx}} \times E \times I$ | $F(4,1688)=9.5$,<br>$p<.001$ | H50 > F50 |
| Hi Sx | R amy. | R STG | 115 | -66.2, 33.8, 16.2 | $P_{\text{HiSx}} \times I$ | $F(2,1688)=12.5$,<br>$p<.001$ | 150 > 50, 100 |

1. STG=Superior temporal gyrus, IOFC=lateral orbitofrontal cortex, gy=gyrus. 2. Significant term from the omnibus model yielding the target region in whole brain analysis. $P_{\text{class}}$ indicates the probability of being in a class, E=Emotion, I=Intensity, and 'x' specifying an interaction. No interactions indicate a main effect. 3. The *post hoc* analysis identifying the contrast yielding the significant term. Simple associations between $P_{\text{class}}$ and connectivity are given for main effects. For interactions, contrasts in $P_{\text{class}}$ and connectivity associations by task condition are indicated by differences in Emotion (A=angry, F=fearful, H=happy) and Intensity (50, 100, and 150%). A description of each interaction is in the main text.

**Figure 2.**
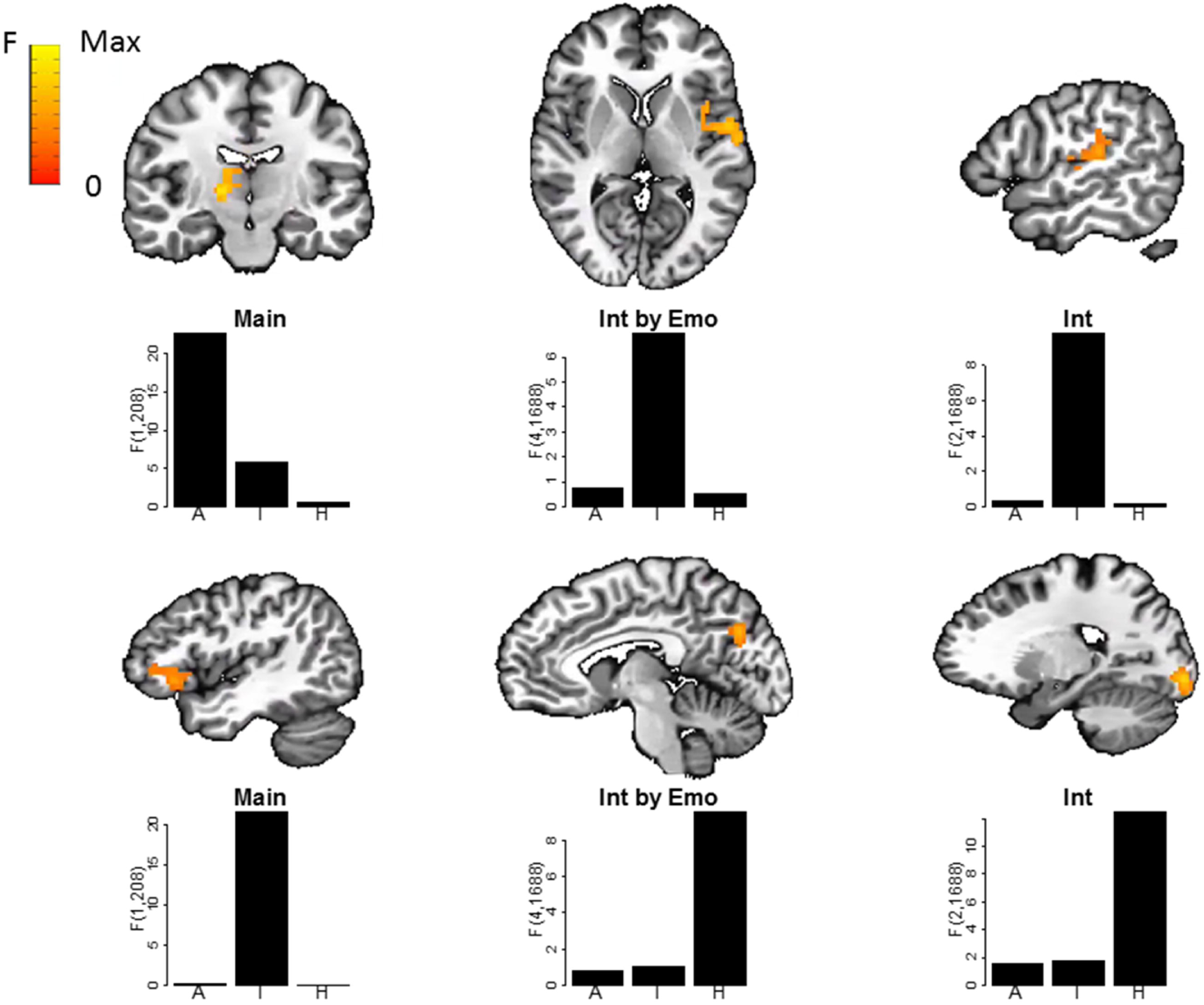
One representative slice of each region of interest at coordinates denoted in Table 2. Below the slices are bar graphs of the relative variance of amygdala connectivity explained by each of the three probabilities of being in an LPA class. From left to right on the x axis the class probabilities are A=Anx/Dep, I=Irr/ADHD, and H=HiSx. The term with which the class probabilities interact is denoted by the title of each bar graph, with Int=Intensity, Emo=Emotion, and Main=the main effect of the class probability. The y-axis represents the terms’ F value, or ratio of variance explained by the term to noise. Because they have the same degrees of freedom, each bar may be interpreted as the connectivity explained by each of the class probabilities relative to each other. Overall, these graphs demonstrate that task-modulated amygdala connectivity is best accounted for by a specific class probability.

Specifically, P_Irr/ADHD_ interacted with emotion and intensity to influence left amygdala connectivity to a region including the left insula and superior temporal gyrus (F_4,1688_=6.96, p<.001). In this interaction, increasing P_Irr/ADHD_ was associated with decreased connectivity in response to 150% fearful and 150% angry faces, both relative to 150% happy (all p’s <.001). P_Irr/ADHD_ also interacted with intensity to influence right amygdala connectivity to the bilateral precuneus (F_1,1688_=9.8, p<.001). Across all emotions, increasing P_Irr/ADHD_ was associated with increased amygdala-precuneus at 50% intensity relative to either 100% or 150% intensity (p’s<.02). Finally, there was a main effect of P_Irr/ADHD_ on left amygdala connectivity on the left lateral orbitofrontal cortex (F_1,208_=21.5, p<.001). Specifically, increasing P_Irr/ADHD_ was associated with decreased connectivity across all stimuli (r=-.31).

A main effect of P_Anx/Dep_ on left amygdala to right thalamic connectivity (F_1,208_=22.7, p<.001) was explained by an association between increasing P_Anx/Dep_ and decreased connectivity across all stimuli (r=-.28).

P_HiSx_ interacted with emotion and intensity to influence right amygdala connectivity to a region that included the left lingual and occipital gyri (F_4,1688_=9.5, p<.001). *Post hoc* contrasts showed this interaction was significant at 50% intensity only. At that intensity, increasing P_HiSx_ was associated with increased connectivity in response to happy vs. fearful faces (p <0.001). P_HiSx_ also interacted with intensity to influence right amygdala connectivity to the right superior temporal gyrus (F_2,1688_=12.5, p<.001). Specifically, increasing P_HiSx_ was associated with increased connectivity in response to 150% intensity, vs. either 100% or 150% intensity (p’s<.001).

*Post hoc* analyses assessing the effects of medication for all regions of interest are presented in Table S3. All findings held, except that the P_Irr/ADHD_ by emotion by intensity interaction did not survive the exclusion of individuals taking stimulant medication.

## Discussion

In a large, transdiagnostic clinical sample of youth, we aimed to identify neural correlates of empirically-defined symptom profiles. We found that, under certain conditions of face-emotion processing, specific regional changes in amygdala connectivity are associated with the likelihood of a specific symptom profile. This study is novel in applying task-based functional connectivity to validate a person-centered clustering of symptoms, which extends (Kircanski *et al*, 2017). As discussed below, the likelihood of being in the symptom class characterized by moderate parent irritability and ADHD symptoms (P_Irr/ADHD_) was associated with amygdala connectivity to frontal, temporal, and insular areas often associated with social or emotional functions. The likelihood of being in the high anxiety/depression class (P_Anx/Dep_) was associated with amygdala connectivity in thalamic regions involved in emotion processing.

P_Irr/ADHD_ had several unique associations to neural response to face-emotions. First, P_Irr/ADHD_ was associated with amygdala-insula/superior temporal gyrus connectivity while responding to high-intensity angry or fearful faces, relative to happy faces. This association may represent a specific amygdala circuitry response to social threat in those expressing irritability/ADHD symptoms. Second, P_Irr/ADHD_ was associated specifically with changes in amygdala-precuneus connectivity by intensity of face-emotion. A relevant, earlier finding was that, across face-emotions, emotion intensity influenced amygdala activation in youth with DMDD (Wiggins *et al*, 2016). Like the irritability/ADHD class in this work, DMDD in the prior study was characterized by irritability and, in most patients, ADHD. These two associations with P_Irr/ADHD_ also correspond to a report by Karalunas *et al*, (2014) on an irritable subtype of ADHD. Specifically, the irritable subtype of ADHD in Karalunas *et al*, (2014) was associated with amygdala dysconnectivity to the precuneus and insula, detected in task-free, ‘resting state’ fMRI. Altogether, these studies converge to implicate aberrant amygdala function to clinical presentations involving both irritability and ADHD symptoms. This supports further investigation of the irritable/ADHD phenotype in youth as a valid psychiatric construct with a specific neural basis.

P_Irr/ADHD_ was also associated with amygdala dysconnectivity to the ventrolateral prefrontal cortex across task conditions. In patients with irritability, ventrolateral prefrontal dysfunction has been reported in several experimental contexts, including reversal learning (Adleman et al, 2011), reward processing (Bebko *et al*, 2014), and face-emotion processing (Coccaro et al, 2007). Consistent with this literature, general amygdala dysconnectivity to the ventrolateral prefrontal cortex has been associated with a tendency towards anger in adults (Fulwiler *et al*, 2012).

P_Anx/Dep_ was associated with a task-general decrease in connectivity between the amygdala and the mediodorsal and lateral aspects of the thalamus. The amygdala and thalamus are core limbic regions consistently activated in fMRI studies of emotion (Kober *et al*, 2008); each contain nuclei that mediate emotional processing in human and animal studies (Metzger *et al*, 2013). Consistent with our results, specific amygdala-thalamic connections have been implicated in fear and anxiety, though their extent and functional significance is unknown (Tovote *et al*, 2015).

Thus, convergence with prior work strengthens the conclusion that these circuits are involved specifically in the expression of irritability/ADHD vs. anxiety/depressive symptoms, suggesting an underlying pathophysiology and objective markers of these clinical presentations. This study goes beyond prior work by directly assessing unique associations of brain circuitry with these common symptom presentations.

This study has several limitations. Given its exploratory nature, replication is needed. Small associations between the class probabilities and motion or gender may bias findings, particularly for the probability of having comorbid irritability and ADHD symptoms. However, the associations are acceptable for applying linear models, we took efforts to minimize these potentially confounding effects, and our findings are concordant with the emerging literature. In addition, associations with high parent reported irritability should be interpreted as preliminary given the small fraction of the sample with this presentation. For task effects on amygdala connectivity to the insula and superior temporal gyrus, we cannot disentangle the effects of stimulant use from the probability of having comorbid irritability and ADHD. This finding does highlight a general limitation in clinical research – that it is unethical to withdraw severely ill youth from medication for fMRI research. Future research will need to address the confounding physiologic effects of stimulants and other medications.

We provide early evidence that patterns of amygdala connectivity during face-emotion processing differ among classes of affective symptom presentations in youth. Future research may replicate these findings as well as their diagnostic specificity and relationship to clinical outcomes.

## Data Availability

The data that support the findings of this study are available on request from the senior author M.B. The data are not publicly available due to them containing information that could compromise research participant privacy and consent.

## Acknowledgements

We thank the participants and families as well as the staff of the Emotion and Development Branch and the Scientific and Statistical Computing Core at the NIMH. In particular, we thank Dan Barlow for programming support and Rick Reynolds, MS, for imaging statistics guidance.

